# Planning and adjusting the COVID-19 booster vaccination campaign to reduce disease burden

**DOI:** 10.1101/2024.03.08.24303201

**Authors:** Laura Di Domenico, Yair Goldberg, Vittoria Colizza

## Abstract

As public health policies shifted in 2023 from emergency response to long-term COVID-19 disease management, immunization programs started to face the challenge of formulating routine booster campaigns in a still highly uncertain seasonal behavior of the COVID-19 epidemic. Mathematical models assessing past booster campaigns and integrating knowledge on waning of immunity can help better inform current and future vaccination programs. Focusing on the first booster campaign in the 2021/2022 winter in France, we used a multi-strain age-stratified transmission model to assess the effectiveness of the observed booster vaccination in controlling the succession of Delta, Omicron BA.1 and BA.2 waves. We explored counterfactual scenarios altering the eligibility criteria and inter-dose delay. Our study showed that the success of the immunization program in curtailing the Omicron BA.1 and BA.2 waves was largely dependent on the inclusion of adults among the eligible groups, and was highly sensitive to the inter-dose delay, which was changed over time. Shortening or prolonging this delay, even by only one month, would have required substantial social distancing interventions to curtail the hospitalization peak. Also, the time window for adjusting the delay was very short. Our findings highlight the importance of readiness and adaptation in the formulation of routine booster campaign in the current level of epidemiological uncertainty.

## INTRODUCTION

As vaccine efficacy wanes over time, one key aspect of the COVID-19 booster vaccination policy is to determine the timing of delivery of the vaccination program to maximize its potential benefits. This is generally informed by several evolving factors: the overall population immunity^1–3^, depending on heterogeneous individual history in terms of vaccination, prior infections, and their combined effect, and on its waning level of protection; the characteristics of the circulating variants^4–6^, especially in terms of disease severity, affected populations, and immune escape; and the expected risk of exposure anticipating an upcoming epidemic wave. Spacing out vaccination campaigns while maintaining a good level of protection in the population during a surge of cases^7,8^ is key to reduce the burden of the disease. However, such planning needs to rely on the above elements, which can hardly be anticipated with accuracy and reliability^9^.

The current epidemiology of COVID-19 remains indeed uncertain, preventing the formulation of routine immunization programs while entering into the first post-pandemic^10^ winter of 2023/2024.. Higher viral activity has been registered in the winter season, corresponding to the traditional influenza season. But in 2023, many European countries recommended spring vaccination campaigns for at-risk individuals, in addition to the fall vaccination campaigns coupled with influenza vaccination programs. In April 2023, the European Center for Disease Prevention and Control (ECDC) recommended member countries to prepare for a continued roll-out of COVID-19 vaccines, particularly during the autumn/winter season, targeting vulnerable populations^11^. Despite the lower peaks in hospitalizations and intensive care unit admissions compared to the pandemic period, the 2023/2024 winter currently experiences an important COVID-19 surge in hospitalizations^12^, coupled with influenza and RSV, pressing individuals to wear masks again and authorities to maintain recommendations for vaccination in full season.

Mathematical models of disease transmission can address uncertainty and provide important insights for prevention and control, accounting for the interplay of key evolving factors^7,13–18^. Modeling undertaken at ECDC was used to provide scenarios comparing a fall 2023 vaccination campaign with one coupled with a spring 2023 vaccination campaign, to formulate the recommendations for member countries^11^. However, the emergence of a new variant with higher transmissibility or immune escape properties may significantly change epidemic conditions and hinder the effectiveness of the planned campaign^8,19,20^, so that rapid response and adjustments may be needed.

Assessing past booster campaigns through modeling can provide insights to inform the formulation of routine COVID-19 vaccination programs. Here, we focus on 2021-2022 winter in France, when the Delta wave followed by the emergence of the Omicron variants challenged the planned booster dosing schedule. We used a COVID-19 multi-strain age-stratified transmission model^21,22^ based on the observed vaccine uptake, estimates of vaccine effectiveness, waning and hybrid immunity, and variants characteristics to expose the benefits and the limits of planning and adjusting a booster campaign in an evolving epidemic context.

## RESULTS

### SARS-CoV-2 epidemic in the Delta-Omicron period

We used a stochastic age-stratified multi-strain transmission model with vaccination (see Methods) and fitted it to COVID-19 hospital admission data (Fig. 1c) and SARS-CoV-2 genomic surveillance data (Fig. 1d) to reproduce SARS-CoV-2 spread in France since the start of the pandemic. Age groups included children (0-10 years old), adolescents (11-18 years old), adults (19-64 years old) and seniors (65+ years old). We validated model results with serological estimates and age-stratified hospitalizations (Fig. S6, S7). This analysis focused on the Delta-Omicron phase (September 2021 – May 2022), that includes the period of the booster campaign, the Delta wave, and the Omicron BA.1 and BA.2 waves (Fig. 1).

**Figure 1.**
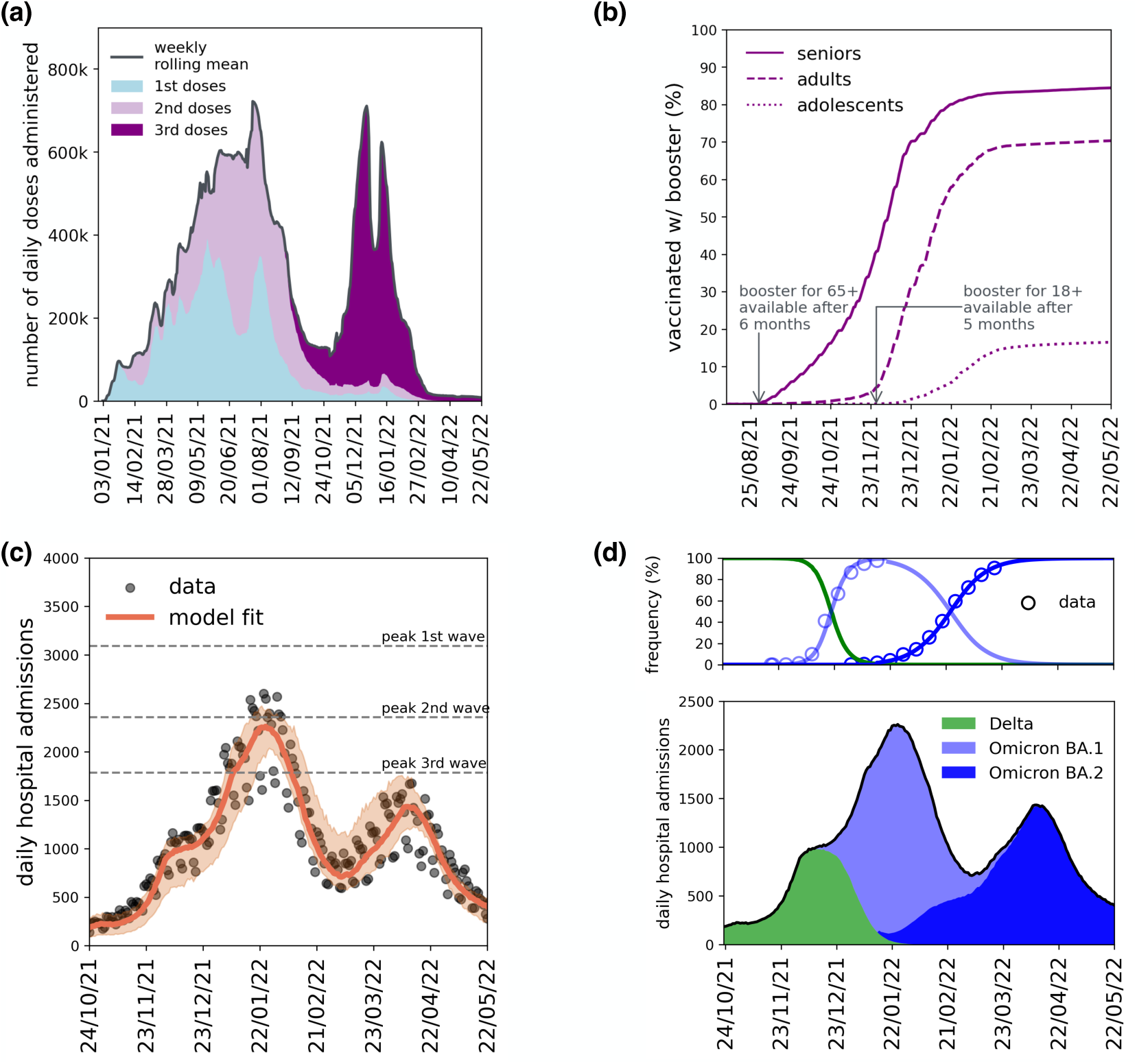
Vaccination campaign and COVID-19 epidemic activity in France during winter-spring 2021-2022. **(a)** Number of daily doses administered in metropolitan France since the start of the vaccination campaign, broken down by first, second and third doses. Panel shows a weekly rolling mean to smooth out weekday/weekend patterns. **(b)** Percentage of individuals who received the 3^rd^ dose (booster) by age class and over time. Line types indicate different age groups, i.e. 65+ (seniors), [19-64] (adults) and [11-18] (adolescents). Arrows indicate relevant dates, i.e. August 31, 2021 when the booster campaign started with eligibility conditioned to 65+ years old and a minimum delay of 6 months since the 2^nd^ dose, and November 27, 2021 when eligibility was extended to individuals with 18+ years and the minimum delay was shortened to 5 months since the 2^nd^ dose. **(c)** Daily number of hospital admissions due to COVID-19 over time. Dots represent the observed data; continuous line and shaded area indicate the output predicted by the model, respectively the median and 95% probability ranges computed over 100 independent stochastic runs. **(d)** Prevalence of Delta and Omicron BA.1 and BA.2 in terms of case frequency (top) and estimated related hospital admissions (bottom). Top: variants’ frequency simulated in the model (continuous lines) and estimated from sequencing data (dots) provided by EMERGEN^24^. Bottom: trajectory of hospital admissions predicted by the model, broken down by variant.

In France, the 2021/2022 winter was characterized by the Delta wave peaking in December 2021, followed by two large epidemic waves, in January and in March respectively, as a result of the spread of Omicron BA.1 and BA.2 (Fig. 1d). The booster campaign started in early September (Fig. 1a), with a gradually increasing coverage in the senior population (Fig. 1b). The booster dose was available for seniors after 6 months from the injection of the 2^nd^ dose, however, the observed rhythm of uptake was initially slower, approximately equal to an effective inter-dose interval of 7 months (Fig. 2a). The vaccination campaign was accelerated in November, in response to the observed increase in epidemic activity due to the Delta variant. First, in mid-November, following changes in the conditions for the validity of the health pass (Methods) and the shortening of the interval for eligibility to 5 months, the uptake rhythm in the senior population increased reaching approximately an effective inter-dose delay of 6 months (Fig. 2a). Then, in late November, booster became available also for the adult population with eligibility conditioned to 5 months since last injection. Consequently, adult vaccination coverage increased until reaching a rhythm of uptake close to an effective inter-dose interval of 6 months (Fig. 2b), i.e. one-month longer with respect to the delay recommended by the policy, similarly to what observed for seniors.

**Figure 2.**
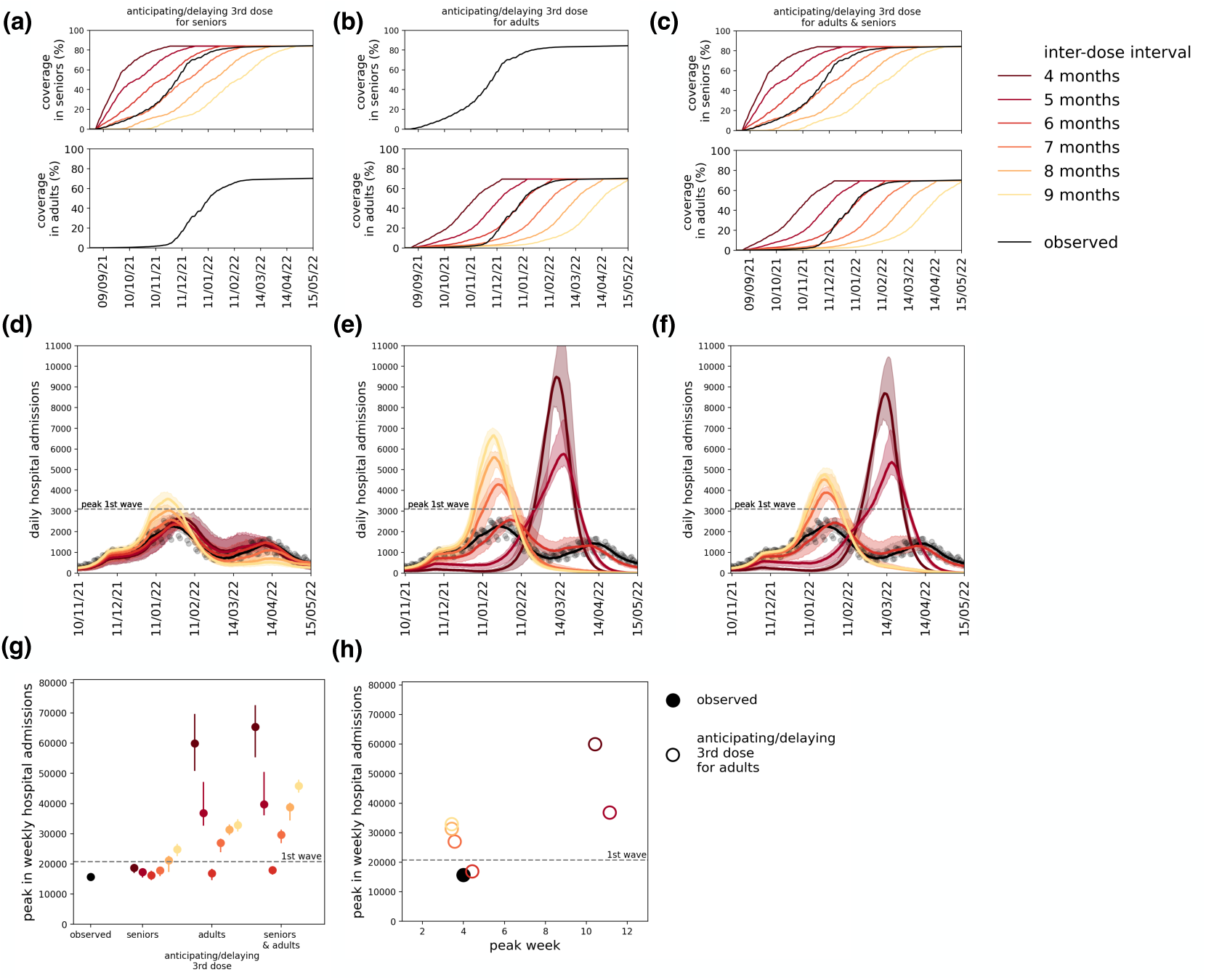
Scenarios with booster campaigns adopting different inter-dose intervals. **(a-c)** Vaccination coverage with the booster dose in seniors (top) and adults (bottom) obtained in three alternative vaccination campaign scenarios anticipating or delaying the 3^rd^ dose for seniors only (left), for adults only (center) and for both age classes (right). Black curves represent the observed vaccination pace; colored lines indicate the vaccination coverage obtained with the booster assuming an effective inter-dose interval, ranging from 4 months (darker color) to 9 months (lighter color). **(d-f)** Epidemic trajectories in terms of daily hospital admissions obtained under the corresponding vaccination scenario shown in panels (a-c). Color code as in panels (a-c); black curve indicates the result of the fit under the observed vaccination campaign. Dots represent the observed data; continuous line and shaded area indicate the output predicted by the model, respectively the median and 95% probability ranges computed over 100 independent stochastic runs. **(g)** Peak in weekly hospital admissions obtained under alternative vaccination campaign scenario anticipating or delaying the 3^rd^ dose for seniors only, for adults only, and for both age classes. Color code as in panels (a-c). The peak registered during the first wave (horizontal grey line) and during the Omicron wave (black dot) are shown for comparison. Symbols and vertical bars indicate median values and associated 95% probability ranges. **(h)** As in panel (g), showing peak in weekly hospital admissions as a function of the week in which the peak occurs, for the scenario anticipating/delaying the administration of the 3^rd^ dose to adults only (see Fig. S8 for scenarios applied to seniors only, and to both seniors and adults).

At the start of December, the first cases associated with the Omicron BA.1 variant were detected in the French territory^23^. The Omicron strain exhibited a significant growth advantage with respect to Delta, and became dominant by the end of December according to genomic surveillance data (Fig. 1d). Consequently, a rapid increase in hospital admissions was observed at the start of January 2022 despite the acceleration in vaccine administration, reaching a peak similar to the one observed during the 2^nd^ wave in winter 2020 in France (approximately 16,000 weekly hospital admissions). Hospitalizations then started to decrease, in absence of additional social distancing measures or evident behavioral changes (e.g. mobility remained stable, Fig. S4), suggesting an effect of building population immunity. Cases and hospitalizations increased again in March 2022, after Omicron BA.2 became dominant, reaching a second peak of hospitalizations (around 10,000 weekly hospital admissions) in April 2022, similar in size to the third wave in spring 2021 due to the Alpha variant. We estimated that, in absence of Omicron BA.2, and assuming the same conditions on contact rates and population mixing, hospitalizations would have continued to decrease up to May 2022 without rebound effects (Fig. S17).

### Impact of vaccine inter-dose delay

To assess the impact of the timing of the booster dose, we explored alternative vaccination scenarios with different effective inter-dose delays between the receipt of the 2^nd^ and the 3^rd^ dose, ranging from 4 months up to 9 months (Fig. 2a-c). To expose the role of different age groups, we considered scenarios in which we varied the inter-dose delay for seniors and adults separately. We found that changing the timing of the booster administration for seniors only, while vaccinating the adult population as observed, would have had limited impact on the trajectory of hospitalizations (Fig. 2d,g). Adult vaccination instead largely impacted the epidemic trajectory (Fig. 2e,f). In the following, we then focused on scenarios where the timing of the booster dose for adults was changed.

After the extension of booster eligibility in late November, requiring an eligibility interval of at least 5 months, the adopted vaccination rhythm for adults reached an effective delay of approximately 6 months, as shown by the similarity of the 6-month delay trajectory with observations (Fig. 2b). Beyond this reference scenario, we identified two epidemic regimes. In the first regime, a longer inter-dose delay (i.e., 7,8,9 months) would have led to a larger epidemic peak in January (27,000 – 33,000 weekly hospital admissions instead of 16,000), because of limited vaccination coverage against BA.1 in December (Fig. 2e). In the second regime, a shorter inter-dose delay (4 or 5 months) would have allowed to suppress the BA.1 wave, however leading to an even larger wave in March due to Omicron BA.2, with hospital peak levels two or three times higher than the peak observed during the first wave in 2020 (Fig. 2e). The considerable waning in vaccine protection in individuals vaccinated too early, e.g., in the scenario with 4-month inter-dose delay, would contribute to the larger epidemic in March. Indeed, Fig. S18 shows that, in absence of waning in vaccine effectiveness, the size of the BA.2 wave would have been halved. Similar results were found if both age classes (adults and seniors) were affected by the change in the inter-dose delay (Fig. 2f,g). The two regimes are clearly visible in Fig. 2h.

### Impact of one-month social distancing

To evaluate and compare the controllability in terms of non-pharmaceutical interventions for the vaccination scenarios with large epidemic waves, we quantified the level of social distancing needed to contain hospitalizations under manageable levels. We modeled social distancing interventions of increasing stringency by reducing contacts rate for a duration of four weeks in the rising phase of the BA.1 or BA.2 wave (Methods), and evaluated the impact in reducing the peak in hospital admissions. We identified the level of *sufficient social distancing* (Methods), i.e., the minimal level of reduction in social mixing, in order to control the epidemic using as a reference the peak observed during the first wave in 2020 (around 21,000 weekly hospital admissions). As before, we focused on the vaccination scenarios anticipating or delaying the 3^rd^ dose for adults only, while seniors are vaccinated with the observed rhythm.

In the first regime, for an effective 8-month inter-dose delay (Fig. 3a), the larger BA.1 wave in January caused by limited vaccination would have been controlled by a sufficient 15% reduction in social mixing starting at the end of Christmas holidays, lowering the peak from 31,000 to 20,000 weekly hospital admissions (Fig. 3c), while avoiding rebound effects in March during the Omicron BA.2 wave. Stronger measures would have postponed the BA.2 peak to late March, however such a peak would have been larger if suppression of Omicron BA.1 was too strict (e.g. 37,000 peak in hospital admissions at the end of March in case of 40% contact reduction in January, Fig. 3c) because of rebound effects due to limited natural immunity. In case of vaccination campaigns adopting an inter-dose delay of 7 or 9 months, sufficient social distancing was found to be at 10% or 20% reduction respectively (Fig. 3d).

**Figure 3.**
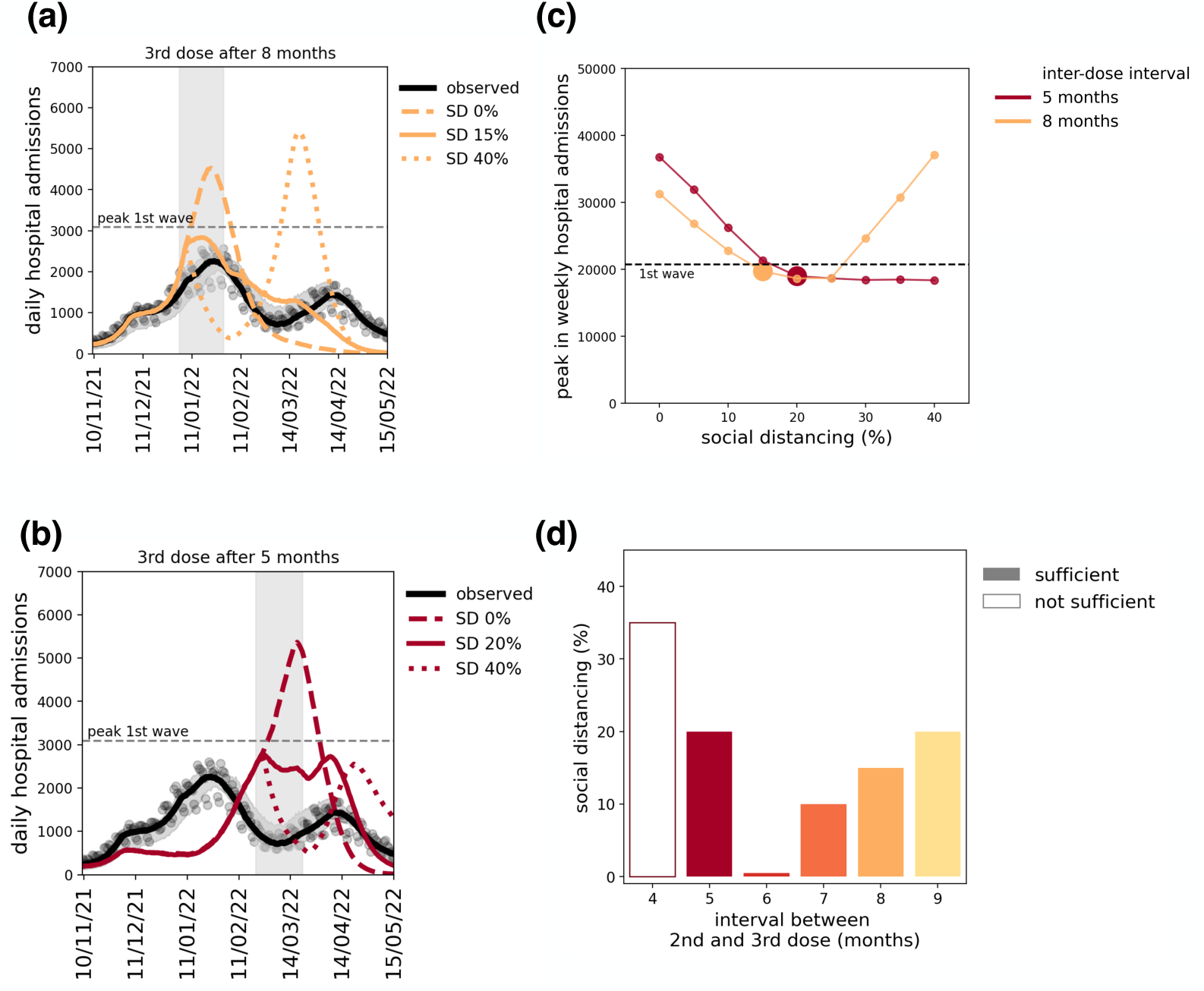
Impact of one-month social distancing. All panels refer to a vaccination scenario anticipating or delaying the 3^rd^ dose for adults only, while seniors are vaccinated as observed. **(a)** Epidemic trajectories obtained with an effective inter-dose interval of 8 months, assuming no social distancing intervention (dashed line) or a one-month measure reducing contact rates by 15% (*sufficient social distancing,* continuous line) or 40% (dotted line) implemented at the end of Christmas holidays (week 1). Vertical shaded area indicates the period of implementation of the measure. Observed data and fitted trajectory are shown in black for comparison. **(b)** As in panel (a), assuming an inter-dose interval of 5 months, with no social distancing intervention (dashed line) or a one-month measure reducing contact rates by 20% (*sufficient social distancing,* continuous line) or 40% (dotted line). One-month social distancing implemented at the end of winter school holidays (week 8). **(c)** Peak in weekly hospital admissions as a function of the level of social distancing implemented. The peak value represents the highest peak among the BA.1 and the BA.2 waves. Horizontal dashed line indicates the peak observed during the first wave, used as a threshold to identify the level of *sufficient social distancing* (larger dots) that allows to keep the epidemic (both BA.1 and BA.2) under a manageable level. Results of panels (a-c) for other inter-dose delays (4,6,7,9 months) are shown in Fig. S9. **(d)** Sufficient social distancing as a function of the inter-dose interval, ranging from 4 months up to 9 months. For the scenarios for which sufficient social distancing was not found (i.e. there wasn’t any contact reduction able to bring the peak below the chosen threshold), the plot shows the level of social distancing leading to the lowest peak possible (labeled as *not sufficient*, void bar).

In the second regime, for an effective 5-month inter-dose delay (Fig. 3b), the higher BA.2 peak in March due to vaccine immunity waning would have required at least 20% reduction of contacts for one month at the end of February, to keep the peak of hospitalizations below the threshold of the first wave (Fig. 3c). In case of a vaccination campaign with a 4-month interval, among the interventions considered, the best-case scenario would have been reached with a 35% contact reduction (Fig. 3d), reducing the peak from 60,000 to 27,000 weekly hospital admissions, still 30% higher than the threshold (Fig. S9). The level of sufficient social distancing was found to be more stringent the higher the hospital incidence at the start of the measure (in other words, the later it is implemented during the rising phase of the epidemic wave, Fig. S11 and Fig. S12), and the higher the reproductive number (Fig. S10). Results do not vary considerably if we apply social distancing for a duration of 3 or 5 weeks (Fig. S19).

### Impact of an adaptive vaccination campaign

To evaluate the potential benefit of an adaptive vaccination campaign, we tested the effect of accelerating vaccine administration in response to the unfolding of the epidemic, in absence of social distancing interventions (Fig. 4). We modeled vaccine acceleration by shortening the effective inter-dose delay on a given week after the start of the campaign (Methods). The observed vaccination campaign in France made booster available for the adult population starting from November 27 (end of week 47 of 2021). The observed rhythm approximately corresponded to a scenario accelerating vaccination in week 48 by shortening the effective inter-dose delay from 8 months to 6 months (Fig. 4a), i.e. a change in eligibility from 7 to 5 months of inter-dose delay, given the observed 1-month difference between eligibility and uptake. A similar outcome was obtained assuming that the initial inter-dose delay was effectively 7 months (i.e. 6-month eligibility, Fig. S13), corresponding to opening vaccination for both seniors and adults at the start of September and accelerating at the end of November.

**Figure 4.**
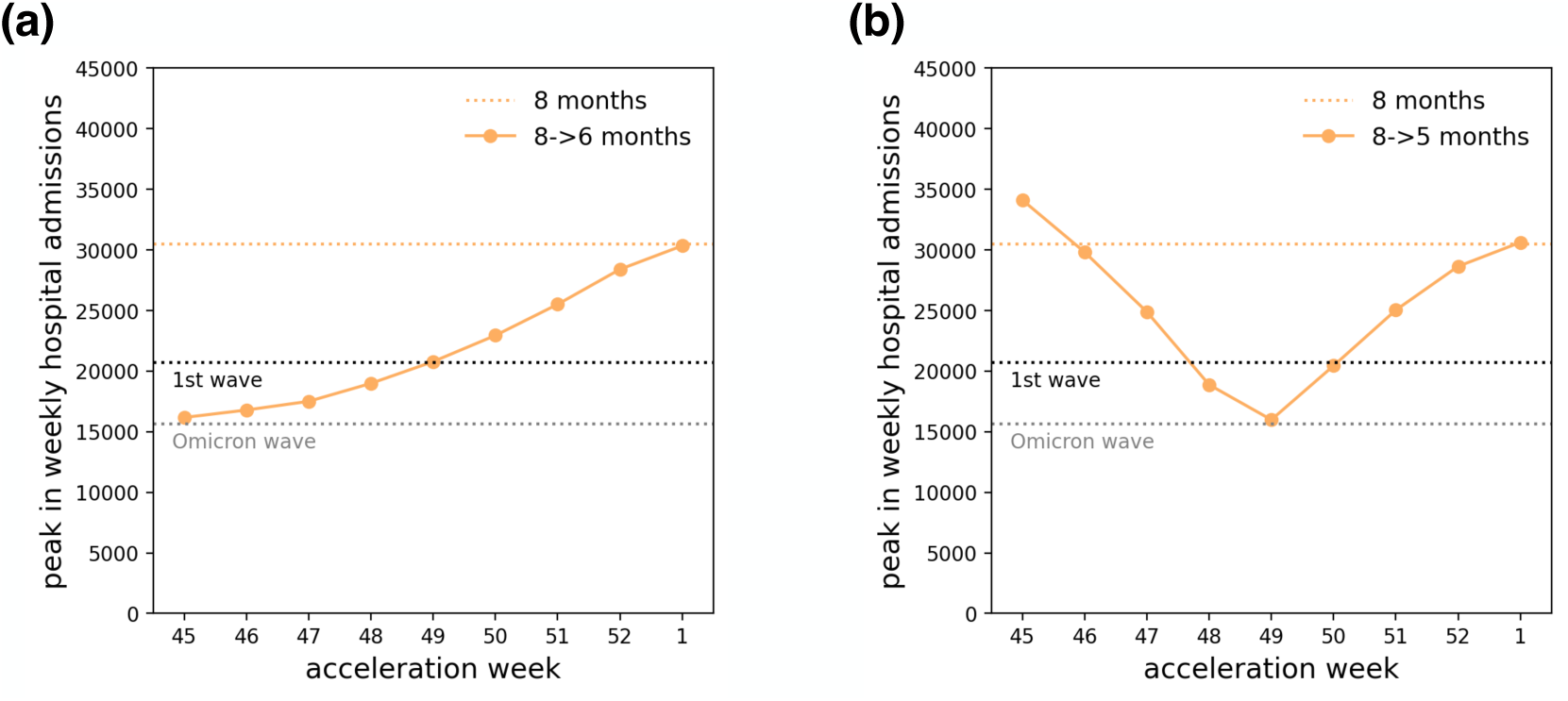
Impact of an accelerated booster campaign. **(a-b)** Peak in weekly hospital admissions (y axis) obtained under a vaccination scenario assuming an inter-dose interval of 8 months shortened to 6 months **(a)** or 5 months **(b)** on a given week (x axis). Colored dotted line indicates the peak obtained in case of no acceleration in the vaccination campaign; gray dotted line indicates the peak level of the observed Omicron wave; black dotted line indicates the peak of the first wave, used as threshold for identifying the level of sufficient social distancing. The peak value represents the highest peak among the BA.1 and the BA.2 waves.

By testing different weeks for the acceleration of the vaccination rhythm, we found that the change in policy should have happened at the latest in week 49 (December 6, 2021), in order to keep the hospitalization peak below the threshold (Fig. 4a), leaving a short period to take action, i.e. approximately 2 weeks after the classification of Omicron as variant of concern (November 26, 2021)^25^. Further shortening the inter-dose delay (from 8 to 5 months effective inter-dose delay) would have reduced the time window for the change of policy: if too early (before week 48, Fig. 4b), the BA.2 wave would not have been controlled, because of the waning of the immunity; if too late (after week 50), vaccination would not have been enough to reduce the hospital burden during the BA.1 wave.

## DISCUSSION

Using a multi-strain age-stratified transmission model accounting for waning in vaccine-induced and natural immunity, we showed that the booster timing adopted in France in preparation for the Delta winter wave in 2021 played a key role in shaping the epidemic in the following months, hitting a delicate balance allowing to control both the Omicron BA.1 and BA.2 waves in January and in March 2022 without requiring additional social distancing. The extension of the eligibility to adults in the general population was key for the control of the successive waves experienced in that winter. The time window for the adjustment of the vaccination program was found to be rather short, of the order of two weeks, highlighting the need for rapid response and readiness.

Different decisions on the timing, anticipating or delaying the start of the booster administration, would have resulted into two different regimes, likely requiring extensive non-pharmaceutical interventions in addition to immunization policies. Not shortening the inter-dose delay or further delaying the booster eligibility would have led to a significant increase in hospitalizations in January 2022 due to Omicron BA.1, requiring a reduction in contact rates of at least 15% for one month to control the peak in hospitalizations. This would have been the result of a late vaccination campaign with respect to the timing of the upcoming wave, therefore not providing sufficient protection to reduce the disease burden in the population. On the other hand, adopting a shorter inter-dose interval would have allowed to fully control the BA.1 wave, at the expense of a larger BA.2 wave later in March 2022, due to waning in vaccine efficacy. The waning in vaccine protection against Omicron infection^26^ resulted to be strong enough to produce the BA.2 wave, despite the level of protection against severe disease and hospitalization remained high^27^. Such a wave would have required stronger measures to be contained (at least 20% reduction in contacts) due to the transmission advantage of Omicron BA.2 over BA.1 (see the Supplementary Material). Even after a large booster campaign, the strong waning in vaccine protection combined with lack of recent natural immunity could rapidly deteriorate the level of immunity in the population, making it vulnerable to potentially new emerging variants. Two years after the emergence of Omicron, this time horizon is now expected to be longer, given the availability of bivalent vaccines^2,28,29^ (targeted on Omicron BA.4 and BA.5), and a large fraction of the population with experience of a past Omicron infection.

All analyses showed that a large booster coverage extended beyond high-risk groups, with the inclusion of the general adult population, was essential to manage the winter waves, similarly to the findings from other countries^30,31^. Despite their reduced risk of developing severe disease with respect to older age classes^32^, adults represent a large fraction of the population having high social mixing, therefore they have the potential to contribute importantly to transmission, generating new cases and ultimately increasing the hospital burden overall. In the current context of large viral circulation but lower hospital admissions compared to the first Omicron waves, thanks to widespread population immunity, the WHO recommends booster vaccination for high-risk groups only, in a base-case scenario where the virus continues to evolve but does not become more virulent^33^. Adapting to scenarios where a new emerging variant shows substantial immune escape or transmission advantage^17,34^ may require revising this policy and extending the vaccination to a larger portion of the population for increased protection. To this end, continuous monitoring of circulating variants through genomic surveillance remains essential^35,36^. Our case study, however, shows that the time horizon for action could be quite short and it would require sufficient logistics and resources to rapidly accelerate the delivery of the booster.

Other countries followed similar steps in the adaptation of the booster vaccination program in late 2021. In the UK, the interval between the second dose and the booster vaccine was reduced from 6 to 3 months after the emergence of Omicron^37^. In mid-November, prior to Omicron, Italy anticipated the date for extension of the age groups eligible for a booster dose with respect to the original plan, and the recommended inter-dose delay was shortened from 6 to 5 months^38^, similarly to France. Israel was the first country to administer the booster dose, with a campaign opening at the end of July, vaccinating up to 80% of the eligible population (16+ years old) by the end of December 2021^30^, allowing the control of the Delta wave without the need to implement strict social distancing. The epidemic trajectory and first booster campaign in Israel are similar to the scenarios explored here with 4 or 5 months delay, with similar results on the importance of the booster rollout timing^30^. The country also administered a second booster dose, during the BA.1 and BA.2 Omicron period, which we do not consider here.

Uptake during the first booster campaign was rather quick and high. In France, we found that the average inter-dose delay was one month longer than allowed by recommendations. This may result from a combination of individual delay in reserving the vaccination appointment and limited availability of doses or administration slots^39^. 70% of adults and 85% of seniors had received the booster by May 2022, likely also prompted by the health pass^40^, however reaching a lower coverage than during the primary vaccination cycle (87% and 92% respectively). Willingness to get a booster dose may further lower due to the evolving epidemic context. A survey conducted in France in summer 2022 found that about 3 out of 4 individuals were willing to get a booster dose for the following winter^41^. However, only 9% of adults and 49% of seniors in France received a second booster dose by May 2023^42^, despite recommendations for one or two shots per year respectively^43^. In the ongoing 2023/2024 winter, only 30% of seniors have received an additional booster dose as of February 4, 2024^44^. Effective communication and recommendations incentivizing vaccination are needed to overcome vaccine fatigue^45^, maximize booster coverage, and limit the significant hospitalizations that COVID-19 continues to cause.

Our work has a set of limitations. First, alternative vaccination scenarios are built by varying the vaccination rhythm and assuming the same conditions of social contacts and intrinsic transmission rate as estimated by fitting the model to the observed epidemic. We did not model changes in individual preventive behaviors in response to a rising epidemic. Survey data for France show that percentage of participants declaring to wear a mask in public places^46^ increased slightly (from 66% to 73%) in the period between October 2021 and January 2022, i.e. in the rising phase of the Delta-Omicron BA.1 winter wave, and then decreased in February after the peak in hospitalizations. In addition, no interventions on social mixing were in place during that period. Second, we did not vary the timing of emergence of Omicron BA.1 and BA.2 across vaccination scenarios, although the level of immunity at the time of the variant’s importation in the country may alter its probability of extinction and affect the moment in which the variant starts to spread steadily in the population. In case the variant emerged at a different moment when population immunity was lower, we expect that our scenarios would require additional social distancing to control the wave. Third, we used an age-stratified transmission model at the national level for mainland France, following prior works^13,47^, therefore neglecting local factors. These include spatial differences in variants’ penetration^48,49^ and the application of social distancing restrictions in our scenario analyses. We note however that non-pharmaceutical interventions were always applied nationwide in France, with limited exceptions^50^.

This study highlights the importance of the timing of the booster rollout to limit COVID-19 disease burden resulting from limited immune protection, due to waning effects or the emergence of newly circulating variants with immune escape properties. As we approach a new phase likely characterized by recurrent seasonal waves, our findings can help strategize future routine vaccination programs, where readiness and adaptation remain key.

## METHODS

### Transmission model

We used a stochastic age-structured two-strain transmission model with vaccination, parameterized using French data on demography^51^, age profile^51^, social contacts^52^, mobility^53^, adoption of preventive measures , and vaccine uptake^42^. We considered four age classes: [0-10], [11-18], [19-64], and 65+ years old, referred as children, adolescents, adults, and seniors respectively. Transmission dynamics follows a compartmental scheme illustrated in Fig. S1 and adopted in previous works^13,22,54,55^, which accounts for latency period, pre-symptomatic transmission, asymptomatic and symptomatic infections with different degrees of severity, and individuals affected by severe symptoms requiring hospitalization. Model details, parameter values and sources are reported in Table S1 in the Supplementary Material. The model is further stratified by vaccine dose, to build vaccine coverage in the population over time according to data on vaccine doses administered in France^42^ (Fig. 1a), and time since vaccination, to model steps of waning in vaccine effectiveness. The model accounts for possible re-infection with Omicron after a prior Omicron or non-Omicron infection, with possible waning in protection against re-infection. Contact matrices describing mixing among age classes are parameterized over time to model change in behavior due to interventions (Fig. S3, S4). Based on pre-pandemic contact data^52^, we built synthetic contact matrices by reducing contact rates for specific locations and age classes, according to mobility data related to workplaces^53^, calendar of school closures^56,57^, and survey data on avoidance of physical contacts^46^, as described in previous works^13,22^.

### SARS-CoV-2 variants

Variant-dependent parameters include the generation time, the transmission advantage and the infection-hospitalization ratio (Section 2 in the Supplementary Material). The model reproduces the co-circulation of two strains, and was applied to describe the Wuhan-Alpha period (February 2020 – May 2021), the Alpha-Delta period (June 2021 – August 2021), and the Delta-Omicron period (September 2021 – May 2022). For the Delta-Omicron period, we modeled the take-over of the sub-lineage BA.2 over BA.1 implicitly by modulating the transmission rate per contact based on the estimated transmission advantage and the proportion of BA.2 over time (Section 6 in the Supplementary Material).

### Model calibration

The model is fitted to hospital admission data since the start of the pandemic (February 2020) up to May 22, 2022, building natural immunity in the population. We used a maximum likelihood approach to fit a step-wise transmission rate. More specifically, in the pre-lockdown phase (February – March 2020), we fitted the starting date of the epidemic and the baseline transmission rate per contact 𝛽_!"#$%&_. Then, we fitted a scaling factor 𝛼_!’()#_of the transmission rate in subsequent time-windows, each one representing a different pandemic phase, based on epidemic activity, behavior and interventions implemented (e.g. pre-lockdown, lockdown, exit phase, summer, curfew). As variant’s transmissibility and effect of interventions are explicitly modeled through the transmission advantage and the synthetic contact matrices, the parameter 𝛼_!’()#_ is meant to absorb other factors potentially affecting the transmission, e.g. mask usage or outdoor/indoor activity. We also fitted the transmission advantage of BA.1 and BA.2 to genomic surveillance data using a binomial likelihood. Details on the inference framework can be found in Section 6 of the Supplementary Material.

### Model validation

We validated the model in terms of percentage of antibody-positive population (Supplementary Material). We modeled the time for seroconversion and seroreversion after infection assuming an exponential distribution with an average of 14 days^58^ and 420 days^59^ respectively. We compared the estimated fraction of antibody-positive predicted by the model with serological estimates^60^ (Fig. S6). We also validated model trajectories by age group with age-stratified hospital data (Fig. S7).

### Infection-induced (natural) immunity

In the pre-Omicron phase (March 2020 – August 2021), we considered full protection against re-infection with the same variant or across variants, given the limited number of re-infections observed in France in this time period^61^, and the estimated highly reduced risk of re-infections^62^. We considered possible re-infection with the Omicron strain, for individuals with a previous Omicron or non-Omicron infection. We did not consider re-infection with a non-Omicron strain after a prior Omicron infection, due to the fast takeover of Omicron sub-lineages over the previous variants. Infection-induced immunity against Omicron infection for individuals with a non-Omicron past infection was set to 62%, following Ref.^63^. Individuals with a prior Omicron infection were assumed to have a higher protection against infection with an Omicron sub-lineage, set at 90%, as suggested in Ref.^64^. Infection-induced protection against hospitalization was set at 88% in case of a prior non-Omicron infection, following Ref.^63^, and assumed to be 95% in case of a prior Omicron infection. Individuals infected twice were assumed to have a reduced infectiousness with respect to a first infection, with a reduction of 50%. We considered waning in infection-induced immunity conferred by non-Omicron infection against Omicron infection, decreasing from 62% to 30% after 6 months according to evidence in Refs.^65,66^ . We assumed no decay for infection-induced protection against hospitalization in the period under study, in line with Ref.^67^ . Parameter values are summarized in Table S3.

### Vaccination and hybrid immunity

We considered vaccination to have an effect against infection, symptomatic infection, hospitalization, and transmission. We used variant-specific estimates of vaccine effectiveness against these outcomes, summarized in Table S4, accounting for evidence of waning in vaccine-induced immunity^26^. We modeled waning in vaccine efficacy after a 2^nd^ or a 3^rd^ dose with step-wise decreasing levels of protection, based on the time since injection. We considered 4 steps of waning, i.e. efficacy within 5 weeks, 10 weeks, 15 weeks and afterwards, by adding layers to the compartmental model. Individuals move between layers based on the time since their last injection or when they receive a new dose. The number of new doses is assigned proportionally to the susceptible and recovered compartment (i.e. not depending on the infection history), according to the vaccination rhythm over time by age class observed in France. We assumed that the 3^rd^ dose is given with priority to the individuals who received the 2^nd^ dose less recently (i.e. 3^rd^ dose is assigned to individuals in the compartment who received the 2^nd^ dose since 15+ weeks). Multiple studies have highlighted that vaccinated individuals with a prior infection have a stronger protection against reinfection with respect to vaccinated-only or infected-only individuals^65,68^. For this reason, we modeled hybrid immunity against infection accounting for the role of prior infection in boosting the protection conferred by the vaccine only. Details are provided in Section 3 in the Supplementary Material.

### Booster campaign in France

The booster campaign in France began at the start of September 2021^69^, with administration of a 3^rd^ dose to seniors (older than 65 years of age) and vulnerable individuals, with a minimum delay of 6 months after injection of the 2^nd^ dose^70^, following recommendations provided at the end of August by the French public health agency Haute Autorité de Santé (HAS)^70^. Starting November 27, 2021, the booster dose became available for the general adult population (18 years old or older)^71^, and the recommended inter-dose interval was shortened to 5 months since the receipt of the 2^nd^ dose. The decision of accelerating the booster campaign relied on the recommendation of the Haute Autorité de Santé based on the observed increase in case incidence in early November due to Delta^72^, prior to the detection of Omicron in the country^23^. The announcement of extending vaccine eligibility came on November 25, 2021^71^, just before the declaration of Omicron as variant of concern by WHO^25^, after the alert from South Africa about rapidly worsening epidemic indicators since mid-November associated with a new SARS-CoV-2 lineage^73^. Booster for adolescents became available starting from January 24, 2022^74^. There were no generalized social distancing restrictions implemented during the booster campaign.

Instead, authorities imposed the health pass as preventive measure. The health pass was an individual certificate required to attend public places (such as bars, restaurants, museums, cinemas, sports centers, cinemas, etc.)^75^. The health pass could be obtained either after a two-dose vaccination scheme (vaccination pass), or, in absence of vaccination, with a negative RT-PCR performed in the latest 72 hours. On November 9, 2021, to incentivize booster uptake in the senior population, French authorities announced additional constraints to extend the validity of the health pass beyond December 15, that required the receipt of a 3^rd^ dose or a negative RT-PCR in the latest 24 hours. Same conditions were applied for the adult population when booster became available, to extend the validity of the heath pass beyond January 15, 2022.

### Scenarios of booster campaigns

We implemented different booster strategies by varying the rhythm of administration of the 3^rd^ dose. More precisely, we focused on the interval between the receipt of the 2^nd^ and the 3^rd^ dose, as this is the reference quantity that is usually set by public health authorities during a vaccination campaign. We tested booster strategies that distribute the 3^rd^ dose with the same rhythm observed for the 2^nd^ dose, with a fixed delay between 4 months up to 9 months since the 2^nd^ vaccination campaign. This delay represents the effective interval between the 2^nd^ and the 3^rd^ dose, and does not necessarily coincide with the interval envisioned by the policy. We considered vaccination scenarios administering the 3^rd^ dose up to the coverage observed during the realized booster campaign (70% for adults and 84% for seniors). Scenarios in which all individuals with a 2^nd^ dose receive a booster are included in a sensitivity analysis (Fig. S15). We considered booster campaigns starting from September 1, 2021, in the main analysis, and tested a starting date on August 1, 2021, in a sensitivity analysis (Fig. S16). In case of short delays, the excess of individuals promptly eligible for booster at the start of the vaccination campaign is uniformly distributed within the following month. To assess the role of different age groups, for any given delay we tested three possible scenarios where this delay is applied the booster rhythm of (i) the senior age class only, distributing the 3^rd^ dose to adults as observed, (ii) the adult age class only, distributing the 3^rd^ dose to seniors as observed, and (iii) both age classes. In all scenarios, we kept the rhythm of administration of 3^rd^ dose for adolescents as observed. We compared scenarios in terms of size and timing of the weekly peak of hospital admissions as a measure of healthcare burden.

### Effect of social distancing

Based on the conditions for contacts and transmissibility fitted on the observed epidemic, we modeled the effect of a one-month social distancing intervention by reducing contact rates by a given percentage, ranging between 5% up to 40%. The starting date of social distancing was fixed based on the French calendar at the start of the week after the end of school holidays (either after Christmas holidays or after the winter break in February depending on the timing of the wave in the scenario under consideration). In a secondary analysis, we varied the date of the implementation of social distancing (Fig. 4, Fig. S11, 12). For each vaccination campaign scenario where we implemented social distancing, we identified the *sufficient* social distancing, i.e. the minimal level of social distancing necessary to obtain a manageable wave, both during Omicron BA.1 and during Omicron BA.2. A manageable wave was defined as an epidemic with a weekly peak of hospitalizations below a threshold, set at the level registered during the first wave (i.e. the highest peak registered in France, approximately 21,000 weekly hospitalizations). In case there exists no reduction in contacts able to keep both epidemic peaks under manageable levels, we consider the most effective social distancing, i.e. the level of social distancing resulting in the lowest peak possible (and we indicate it as *not sufficient* social distancing). We test the effect of a one-time intervention; scenarios with a large epidemic rebound after the lifting of social distancing should not be considered to be realistic, as they would require the implementation of a second measure. We fixed the duration of social distancing interventions at 4 weeks, and we tested different durations in the sensitivity analysis (Fig. S19).

### Acceleration of the booster campaign

We tested interventions aimed at reducing the epidemic activity by accelerating the rhythm of vaccination, as an alternative to implementation of social distancing. We modeled this in terms of effective delays between the 2^nd^ and the 3^rd^ dose. Given a scenario of booster campaign with an initial interval of 7 or 8 months, we simulated scenarios in which the interval of administration is shortened to 5 and 6 months starting from a given week. The excess of individuals who become eligible for the booster when the inter-dose delay is shortened are absorbed within two months by distributing uniformly their number in the period. The duration of two months is chosen so that the vaccination rates remain feasible and do not largely exceed the maximum daily number of administered doses registered during the actual campaign in France (Fig. S14).

## Data Availability

Data are available online at cited references.

## ACKNOWLEDGMENTS

This study was partially funded by: Agence Nationale de la Recherche project DATAREDUX (ANR-19-CE46-0008-03) to VC; ANRS–Maladies Infectieuses Émergentes project EMERGEN (ANRS0151) to VC; Horizon Europe grant ESCAPE (101095619) to VC; EU Horizon 2020 grant MOOD (H2020-874850, paper 105) to VC. The study is catalogued as MOOD105. The contents of this publication are the sole responsibility of the authors and don’t necessarily reflect the views of the European Commission.

